# Protocol for: Association of antipsychotic medication usage and employment outcomes in patients with schizophrenia spectrum disorders: A Danish register-based study

**DOI:** 10.1101/2025.10.02.25337161

**Authors:** Ricardo Twumasi, Maximin Lange, Carsten Hjorthøj, Oliver Howes, Frederikke Hørdam Gronemann, Merete Nordentoft, Merete Osler

**Affiliations:** Copenhagen Research Center for Mental Health – CORE, Mental Health Centre Copenhagen, Copenhagen, Denmark; Section of Epidemiology, Department of Public Health, University of Copenhagen, Copenhagen, Denmark; Department of Psychosis Studies, Institute of Psychiatry, Psychology & Neuroscience, King’s College London, London, UK; South London and Maudsley NHS Foundation Trust, London, UK; Center for Clinical Research and Prevention, Bispebjerg and Frederiksberg Hospitals, Copenhagen, Denmark

## Abstract

**Background:** Schizophrenia spectrum disorders are associated with higher risk of unemployment for the individual. Antipsychotic usage is linked to less risk of relapse, fewer hospitalisations and lower rates of mortality. This study investigates the use and discontinuation of antipsychotics in individuals with psychosis in relation to employment and other functional outcomes.

**Methods:** Using Danish nationwide registers, we identify individuals aged ≥18 with first-time nonaffective psychosis diagnosis (ICD-10 F20-F29) during 1995-2024. Antipsychotic exposure is categorised as continuous use (≥80% coverage), intermittent use (20-80%), sustained discontinuation (<20%), or no use. Functional outcomes include: employment status, benefit code, hospital readmission. Within-subject analyses use each individual as their own control, with time-varying covariates and 6-, 12-, and 24-month lagged effects. Models adjust for treatment sequence, time since diagnosis, and concurrent psychotropic medication use.

**Hypotheses:**

1. For individuals with schizophrenia spectrum disorders, periods of antipsychotic treatment will be associated with higher likelihood of
  a. employment/ education/training and
  b. independent living, relative to periods without such treatment.

*Exploratory hypotheses:* Periods of long acting injectables or clozapine treatment will be associated with higher likelihood of employment/ education and independent living relative to periods with other antipsychotic treatment.

## Methods

### Patients

Included in our analysis will be patients first diagnosed with functional psychosis (ICD-10 F20–F29), aged ≥18 years, from 1995 to 2024 in Denmark. Follow-up ends at death, emigration, or end of data linkage (December 31, 2024), whichever occurs first. Individuals are excluded if emigration, mandatory forensic psychiatric treatment, or death occurs within the first 2 years after diagnosis, as these prevent adequate exposure and outcome assessment during the critical early illness period.

### Databases

Data will be extracted from the Danish National Prescription Register (NPR) (Pottegård et al., 2017), the Befolkningen register (BEF) (Thygesen et al., 2011) Employment Classification Module (AKM) (Petersson et al., 2011), Danish Register for Evaluation of Marginalization (DREAM) (Hjollund et al., 2007), Danish Education Register (UDDA) (Jensen & Rasmussen, 2011), Danish National Patient Registry (Schmidt et al., 2015) and Danish Psychiatric Central Research Register (Mors et al., 2011) and The Danish National Hospital Medicine Register (SMR) (Andersen et al., 2024). These acronyms are based on the Danish names for these databases. The registers will be linked using the unique individual person identification number attached to all Danish citizens.

The NPR serves as the primary data source for antipsychotic exposure assessment, providing comprehensive dispensing data from community pharmacies from 1995-2024. The SMR, established in 2018, will be used for sensitivity analyses to validate findings using hospital prescription data for the period 2018-2024, particularly for inpatients who may not fill prescriptions at community pharmacies.

### Exposures

The primary exposure will be patterns of antipsychotic medication use over time, categorised into use vs non-use. Exploratory further categorisation could be continuous use, intermittent use, sustained discontinuation, and no use.

We will not differentiate between second- and first-generation, or long acting injectables and oral formulations or clozapine. We may differentiate later on, as an exploratory investigation.

When clozapine was received, a 6-month minimum is required (start of prescription being Index point). Antipsychotic use or non-use is thus a time-varying exposure.

The SMR will be used for sub-group analysis, as this database was established in 2018, it only contains hospital prescriptions from 2018-2024.

### Outcomes

The main outcome measure will be start of employment, education or vocational training, regardless of level of compensation or diagnoses.

Secondary outcome measure(s) will be living independently, having children, co-habitation or marriage status, being in receipt of social benefits.

#### Potentially

Type of employment (blue collar vs white collar). Tertiary outcome measures will include:

- Gross annual income (derived from the Income Statistics Register)
- Income from employment specifically (excluding transfer payments)
- Income quartile relative to age-matched general population
- Time to first income after unemployment period

Income data will be inflation-adjusted to 2025 values using Statistics Denmark’s consumer price index.

### Categorisation

*DREAM* information is based on a hierarchy prioritised according to the benefits received in that week, i.e. sickness absence transfer payments rank higher than unemployment transfer payments.

To track labour market transitions, labour market affiliation will be divided into six mutually exclusive categories based on DREAM hierarchical prioritisation:

1. Productive engagement (primary outcome) is defined as person-weeks meeting ANY of the following criteria:
  - Employment: No DREAM benefit code AND monthly employment field category code present
  - Education: DREAM code 224 OR UDDA registration OR SU receipt
  - Vocational training: DREAM codes 133-139 For analytical purposes, productive engagement is coded as a binary time-varying variable assessed weekly.
2. Sickness Absence:
  - Full-time and part-time sickness benefit (sygedagpenge)
  - DREAM codes: 210-213, 220-221
3. Employment-Ready Transfer Payments:
  - Unemployment insurance (a-kasse) benefits
  - Employment-ready cash assistance (arbejdsmarkedsparate kontanthjælp)
  - DREAM codes: 110-113, 130, 133-139
4. Passive Transfer Payments:
  - Not employment-ready cash assistance
  - Social assistance
  - Integration benefits
  - DREAM codes: 140s, 150s, 160s
5. Disability Pension:
  - Permanent disability pension (førtidspension)
  - Early retirement due to disability
  - DREAM codes: 610-619
6. Out Of Labour Force:
  - Death
  - Emigration
  - No benefit and no employment recorded

This categorisation allows for granular analysis while maintaining comparability with previous Danish register studies

### Study Design

This study is a nationwide, register-based cohort study. Index point (cohort entry) is the date of first schizophrenia diagnosis (or discharge from hospital for those who had their first diagnosis recorded in inpatient care). Follow-up ends at death, emigration, or the end of data linkage, whichever occurred first.

### Statistical Analysis

We will report labour market affiliation at baseline (index date) and at 6, 12, and 24 months post-diagnosis using descriptive statistics. The association between AP exposure and employment outcomes will be assessed among individuals who change medication status during follow-up (switchers). Primary analyses will be stratified by baseline employment status (employed vs. unemployed at diagnosis) to examine whether treatment effects differ by initial functional level.

Changes over time in categories will be illustrated in Sankey plots. Benefit distributions will be visualised with violin plots. Individual level trajectories with swimmer plots. Survival and time to event visualisations with Kaplan-Meier curves. Odds ratio’s will be visualised with forest plots.

We use a within-subject design to control for time-invariant confounding factors, acknowledging this restricts our sample to individuals who change exposure status during follow-up (i.e., switch between medication use and non-use). To understand potential selection bias, we will compare characteristics of included patients (those who change exposure) versus excluded patients (those who remain consistently on or off medication). We categorize antipsychotic exposure into continuous use (≥80% medication coverage), no use (0% coverage), intermittent use (20-80% coverage), and sustained discontinuation (<20% coverage over 6+ months).

Time periods of continuous antipsychotic use are compared with periods of intermittent use and non-use within the same individual.

Exposure categories are modelled as time-varying covariates, with individuals potentially contributing person-time to multiple categories during follow-up. The model includes lagged effects to account for delayed impact of medication changes and interaction terms between exposure patterns and time since initiation.

Given the complexity of accurately determining labour market status from Danish register data, we employ a hierarchical algorithm to classify each person-week into analysable vs. non-analysable periods:

### Non-analysable periods (excluded from primary analysis)

#### 1. Ongoing unemployment with active benefit receipt

Continuous receipt (≥4 consecutive weeks) of unemployment-related benefits, specifically:

- Unemployment insurance benefits (DREAM codes 110-113)
- Employment-ready cash assistance (DREAM codes 130, 133-139)
- Passive transfer payments (DREAM codes 140-169)
- Sickness absence benefits (DREAM codes 210-213, 220-221)

The rationale for this is that the outcome (employment/education/training) cannot occur by definition during these states; however, transitions out of these states form the basis of time-to-event analyses.

#### 2. Psychiatric inpatient admissions

This is defined as any week containing ≥1 day of psychiatric hospital admission.

The rationale for this is that employment should not be possible during hospital admission; medication exposure may differ from community prescriptions; labour market status is artificially constrained

#### 3. Indeterminate labour market status

Weeks where exposure status cannot be reliably determined due to:

- Incomplete medication coverage data (e.g., recent immigration, data linkage gaps)
- Transition periods between exposure categories (<14 days in new category)

### Analysable Periods (included in primary analysis)

#### 1. Employment (productive engagement)

Weekly criteria: No DREAM benefit code registered in a particular week.

AND Monthly criteria: Presence of employment field category code (from AKM register or DREAM monthly employment variables) indicating:

- Active employment with income from work
- Registered occupation/industry code
- Employer identification number

Because employment status is determined monthly while DREAM benefits are weekly, we assign employment status to all weeks within a calendar month if the monthly employment field indicates employment.

For edge cases, we will consider that in weeks spanning two calendar months, we will use the employment status from the month containing the majority of days in that week.

Individuals receiving the prison employment code will be retained in analyses but flagged for sensitivity analysis. While the general Danish incarceration rate is low (71 per 100,000), individuals with schizophrenia have elevated criminal justice involvement. Prison-based employment differs qualitatively from community employment, however, obtaining prison registry access requires separate data approval beyond the scope of this protocol.

#### 2. Education/training

- DREAM codes indicating full-time education (code 224) or vocational training (codes 133-139)
- Education register (UDDA) confirmation of active enrolment
- Receipt of SU (Statens Uddannelsesstøtte - student grant) further validates educational activity

#### 3. Disengagement from labour market and benefit systems

Weekly criteria: No DREAM benefit code registered

AND Monthly criteria: No employment field category code (i.e., person is not registered as employed)

AND Duration criteria: Persisting for >4 consecutive weeks

This represents a distinct category indicating withdrawal from both formal employment and the social security system. Such individuals may be:

- Financially supported by family/partners without being registered as employed
- Engaged in informal/undeclared work
- Living abroad without formal emigration registration
- Genuinely disengaged from economic activity

Analytical approach: These periods are flagged separately and analysed in sensitivity analyses to examine whether medication-outcome associations differ when including vs. excluding disengaged periods

Given the almost 25-year observation period, lagged effects are analysed at 6, 12, and 24 months to capture both immediate and long-term relationships between medication changes and employment outcomes. These windows account for potential delayed effects and secular trends in employment patterns over the extended study period.

Time-invariant factors are eliminated by the within-subject design. Thus, adjustments only include temporal order of treatments (sequence of different antipsychotics and non-use, as 1, 2, or 3 vs. >3), time since cohort entry, and time-varying use of other psychotropics.

Concurrent psychotropic medication use includes time-varying indicators for:

- Antidepressants (ATC N06A)
- Mood stabilizers (ATC N03A, N05AN)
- Anxiolytics/hypnotics (ATC N05B, N05C)
- Psychostimulants (ATC N06B)

Coded as binary (yes/no) variables for each class at each time point

We use within-individual analyses, stratified Cox regression models for employment outcomes, mixed-effects survival models for employment duration, multilevel multinomial logistic regression for employment type, and linear mixed models for income outcomes. All models yield adjusted hazard ratios and 95% confidence intervals.

Sensitivity analyses will be conducted in subjects who did not use antipsychotics within one year before cohort entry to ensure that they were truly incident cases, as well as in those without unemployment, education or training at baseline (as previous unemployment increases the chances of having unemployment again), censoring the first 30 days of each exposure (to control the time period of suboptimal effect of antipsychotics), using unemployment only as an outcome, and in time periods of <2 years, 2–5 years, and >5 years after cohort entry. In addition to the main comparison (use, intermittent use, non-use of antipsychotics), we also compare individual antipsychotics. Only unemployment >90 days will be considered for sensitivity analyses, representing longer-term loss of functioning.

Subgroup analyses by sex and baseline education level will also be conducted.

We note that a key methodological challenge is confounding by indication in medication non-use. The no-medication group likely contains two distinct subgroups: those with inherently better prognosis who can maintain employment without medication, and those who would benefit from medication but aren’t taking it. This heterogeneity could create misleading associations between medication non-use and employment outcomes.

Our analytical strategy addresses this through several approaches: First, we use within-subject analyses where each individual serves as their own control, comparing employment during periods of medication use versus non-use. Second, we assess baseline functioning through multiple indicators: employment status at diagnosis, source of first diagnosis (inpatient vs. outpatient care), educational level, and diagnosis type (particularly F23, acute transient psychosis) as stratification variables. Third, we conduct sensitivity analyses in subgroups defined by these baseline characteristics and initial care patterns.

Baseline (Index date - first schizophrenia diagnosis):

- Age, sex, ethnicity, education level, diagnosis (F20-F29 subtype), comorbidities, setting of first diagnosis (inpatient vs outpatient)

Time-Varying (assessed at regular intervals during follow-up):

Every 13 weeks (quarterly):

- Labour market status (DREAM categories)
- Current psychiatric medications
- Residential status (independent vs. institutional)

Every 52 weeks (annually):

- Relationship status (cohabitation/marriage from CPR)
- Educational attainment updates
- Income data

Event-based:

- Hospital admissions and discharges
- Changes in antipsychotic regimen
- Changes in comorbidity status

Sample size should be sufficiently large enough to avoid de-anonymisation. Data will only ever be presented on a group level, and data will be stored in a pseudorandomised state.

Additional sensitivity analyses will include:

- Dose-Response Relationship
- Washout Period Analysis:
- Competing Risks Sensitivity (Fine-Gray subdistribution hazard models)
- Time-Period Stratification
- Propensity Score Matching
- E-Value Calculations (calculating E-values to quantify the minimum strength of association that unmeasured confounders would need to fully explain observed effects)

### Software

Data preparation and analyses will be conducted in SAS, Stata, R and python.

### Protocol adherence and deviations

This protocol will be followed as specified. Any necessary deviations (e.g., due to data availability limitations discovered during analysis) will be:

1. Documented with justification
2. Clearly indicated in the final manuscript
3. Distinguished from pre-specified analyses

Exploratory analyses not specified here will be clearly labelled as post-hoc in all publications.

## Ethics

The study was approved by the Regional Data Protection Agency (P-2020-88). According to Danish law, additional ethical approval is not required for purely register-based studies.

## Data Availability

All data produced in the present work are contained in the manuscript

## Notes

### Competing Interest Statement

The authors have declared no competing interest.

### Funding Statement

This study did not receive any funding

